# Eccentric and Concentric Torque Feedback Training Induce Similar Clinical Improvements but Distinct Triceps Surae Motor Unit Adaptations in Non-Insertional Achilles Tendinopathy: A Randomized Controlled Trial

**DOI:** 10.1101/2025.06.20.25330016

**Authors:** Ignacio Contreras-Hernandez, Deborah Falla, Michail Arvanitidis, Francesco Negro, David Jimenez-Grande, Eduardo Martinez-Valdes

## Abstract

**Background:** Eccentric exercise (ECC) is widely recognized as an effective treatment for non-insertional Achilles tendinopathy (NIAT); however, the mechanisms underlying its apparent superiority over concentric exercise (CON) remain poorly understood. This randomized controlled trial aimed to investigate changes in triceps surae motor unit firing properties, pain, function, and AT morpho-mechanical properties following a 6-week intervention involving torque feedback training with isolated ECC and CON contractions in individuals with NIAT.

**Methods:** Twenty-six individuals with NIAT were randomized to ECC or CON training. Motor unit firing properties (mean discharge rate [MDR], recruitment and de-recruitment thresholds, and torque–firing cross-correlation and neuromechanical delay) were assessed in the medial gastrocnemius (MG), lateral gastrocnemius (LG), and soleus (SO) muscles using high-density surface electromyography (HD-sEMG) during isometric plantarflexion at 10%, 40%, and 70% of maximal voluntary contraction (MVC). Pain, function (VISA-A), and tendon properties (ultrasound, elastography) were measured at baseline, week 3, and week 6.

**Results:** Both groups showed similar improvements in pain (P < 0.0001) and VISA-A scores (P < 0.001). Tendon stiffness increased in both groups by week 3 but was higher in ECC by week 6 (P = 0.02). Motor unit adaptations differed: CON demonstrated an increase in MG MDR at 40% MVC, while ECC showed a decrease (interaction: P = 0.0008). Only ECC led to increased de-recruitment thresholds in the LG at 70% MVC (P < 0.0001). However, both groups exhibited reduced MDR in the LG at high-force levels (P < 0.05). Additionally, both interventions reduced GL torque–firing relationships (P = 0.025) and decreased SO neuromechanical delay (P = 0.031).

**Conclusion:** A 6-week visuo-motor torque feedback training program involving isolated CON or ECC contractions leads to comparable improvements in clinical outcomes. However, contraction-specific and muscle-specific changes in motor unit function and tendon stiffness suggest distinct neuromechanical adaptations. These differences may underlie the observed effects and warrant further investigation in longer-term studies to determine their impact on long-term clinical outcomes.

**HIGHLIGHTS:**

- Changes in clinical outcomes, tendon morphomechanical properties, and triceps surae motor unit behavior were assessed for the first time following a 6-week rehabilitation protocol for non-insertional Achilles tendinopathy, involving isolated concentric (CON) or eccentric (ECC) contractions.
- Both CON and ECC training led to comparable improvements in pain, self-reported outcomes, and perceived function.
- Despite these similar clinical improvements, CON and ECC protocols produced distinct adaptations in estimated tendon stiffness and motor unit firing characteristics.
These neuromechanical differences may indicate potential long-term divergences between training modalities, which should be explored in studies with extended intervention periods.

## INTRODUCTION

Non-insertional Achilles tendinopathy (NIAT) is the most prevalent tendinopathy of the lower extremity ^5^. Albeit our improved understanding of this condition, it remains a slow-progressing injury ^6^, with full recovery taking a year or longer, and its reinjury rate is high ^7, 8^. The etiology of the NIAT remains unclear and it is likely to be multifactorial ^9, 10^. However, from a mechanobiological perspective, repetitive mechanical tissue overuse seems to be a predominant cause ^10, 11^. Thus, excessive loading of the tendon may result in structural damage that, with time, can lead to a degenerated and weakened tendon ^12^. Recent evidence has identified that deficits in the performance of the medial gastrocnemius (MG), lateral gastrocnemius (LG), and soleus (SO) muscles may be a key factor ^13, 14^ since these muscles may influence the magnitude and distribution of the AT load, strain, and stress ^15, 16^.

Morpho-mechanical changes induced by NIAT include an increase in tendon cross-sectional area (CSA), thickness, volume ^18–22^, transverse strain, hysteresis, and a decrease in stiffness and Young’s modulus ^19, 20, 23, 24^. Nevertheless, the impact of different exercise interventions on the morpho-mechanical properties of the Achilles tendon (AT) in individuals with NIAT has shown some ambiguous results. One review focusing on eccentric exercise concluded that tendon thickness does not change in parallel with improved clinical outcomes ^25^. However, a randomized controlled trial comparing heavy-slow resistance exercises and eccentric exercises in individuals with NIAT reported reduced tendon thickness in both groups ^26^. Notably, following an exercise intervention, some tendons may experience an increase in collagen content and size while simultaneously reducing their water content, resulting in unchanged tendon thickness ^25^. Therefore, the AT adaptations to exercise might only be detected by assessing its mechanical properties (i.e., stiffness) ^25^.

The central nervous system regulates muscle forces by modifying the recruitment and discharge rate of the associated motor units ^27^. Recent studies have shown that motor units firing properties can adapt to changes in muscle-tendon properties ^28, 29^. Thus, changes in motor unit firing properties may be behind injury-induced adjustments in motor performance ^27^. It has been suggested that exercise may alter motor unit firing properties, thereby improving the mechanisms of force generation^30^. However, few studies have examined motor unit firing properties in individuals with chronic musculoskeletal conditions, and none have investigated changes in the motor unit firing properties of the triceps surae muscle following an exercise intervention in individuals with NIAT.

Exercise interventions have the highest level of evidence for the management of NIAT ^31^. These interventions aim to provide controlled mechanical loading to the tendon to promote remodeling, decrease pain, and enhance strength of the triceps surae muscle ^6^. Although optimal loading parameters have not been determined, it has been suggested that tendons have a more favorable response to high loads with longer durations ^32^. Historically, exercise interventions based on AT loading have focused on eccentric contractions; however, exercise interventions including concentric contractions or a combination of both have also shown good results^8, 26^. Recently, it has been proposed that isometric exercises may be helpful for the management of the NIAT at early stages^33^; however, there is no evidence supporting its superiority over any other exercise modality^34^. Consequently, the idea that the tendońs response to mechanical load depends on the type of muscle contraction applied has been questioned^32, 35^. Furthermore, there is a lack of studies assessing the effects of pure eccentric or concentric contractions on NIAT since current rehabilitation protocols employed in clinical practice are unable to isolate these contractions^36^. Studies assessing the effect of exercise in individuals with NIAT usually have insufficient control over the load, speed, pain tolerance, or range of movement in which the exercises were performed. Therefore, we have developed an intervention approach based on slow-speed visuo-motor training that uses an isokinetic dynamometer to control tendon loads while providing visual feedback of the exerted torque to the participants. This approach allows for increased time under tension, controlled loading throughout the range of movement, individualized contraction intensity based on each participant’s capacity and pain tolerance to improve motor performance during exercises.

We aimed to investigate the changes in triceps surae motor unit firing properties, pain and function, and AT’s morpho-mechanical properties after applying a 6-week intervention protocol based on either controlled eccentric or concentric contractions in individuals with NIAT. We hypothesized that the intervention protocol based on eccentric contractions will be more effective in inducing changes in triceps surae motor unit firing properties, pain, function, and AT’s morpho-mechanical properties in individuals with NIAT.

## METHODS

Two-arm, parallel-group, randomized controlled trial conducted from October 2021 to April 2023 at a laboratory within the School of Sport, Exercise and Rehabilitation Sciences of the University of Birmingham, UK. This study was approved by the STEM Ethical Review Committee, University of Birmingham, UK (ERN_20-0604A) and registered under ISRCTN registry (ISRCTN46462385). The study was conducted according to the declaration of Helsinki and reported following the CONSORT guidelines ^37^. All participants signed written informed consent prior to participation. The participant flowchart can be seen as ***supplementary figure 1***.

Participants with NIAT attended the laboratory over a six-week period for both experimental and training sessions. Experimental assessments were conducted at baseline (Session 1), after three weeks (Session 2), and after six weeks (Session 3). Training sessions took place 2–3 times per week throughout the intervention. Participants with NIAT were randomly allocated into two groups: eccentric (ECC) or concentric (CON) training. If individuals with NIAT presented bilateral symptoms, the most symptomatic leg was assessed and trained. All participants were instructed to avoid any vigorous physical activity within 24 hours before each experimental session.

Based on power analysis calculations, a total of 26 individuals with NIAT (ECC group= 13, and CON group= 13) were required for this study. This sample size considers a power= 0.80, alpha=0.01, 25% loss of participants and an effect size (d) of 1.7 calculated from the study of Yu et al. ^38^, which investigated the effect of 8 weeks of either eccentric or concentric training on pain levels in individuals with NIAT.

### Participants

Twenty-six participants with NIAT (14 males, 12 females, 29.04 ± 8.46 years, 75.09 ± 17.20 kg, 173 ± 9.16 cm) were recruited. Inclusion criteria included men or women aged 18 to 55 years old (range chosen to minimize the influence of age in tendon properties ^39^). Inclusion criteria were as follows: presence of NIAT determined by an experienced physiotherapist based on clinical findings, physical examination, and ultrasound imaging. Clinical findings encompassed the Victorian Institute of Sports Assessment-Achilles Questionnaire (VISA-A) ^40^, NRS (Numeral Rating Scale) ^41^, and symptoms for more than 3 months ^26^. Individuals with a VISA-A score less than 90 and NRS score ≥ 2 were considered for the study ^42^. Ultrasound imaging was used to assess the echoic pattern and tendon thickness^43^.

The exclusion criteria for the ECC, CON groups were as follows: (1) systemic or inflammatory conditions including malignancy, neuromuscular disorders, and rheumatic diseases, (2) current or history of chronic cardiovascular, neurological, or respiratory diseases, and (3) history of lower limb surgery. Additionally, participants were excluded if they had been diagnosed with insertional Achilles tendinopathy or if they received a corticosteroid injection in the AT in the last 12 months.

### Randomization, allocation, and blinding

Individuals with NIAT were randomized by one researcher (EM-V) in a 1:1 allocation ratio to either ECC or CON groups using a computer-generated simple scheme randomization. EM-V secured the randomization code using password-protected files. EM-V released the randomization code once each participant with NIAT completed the first experimental session. To achieve treatment blinding, a third researcher (M-A) encoded the participants ID using blindr software (https://github.com/U8NWXD/blindr). IC-H performed experimental and training sessions, however, did not have access to the participant’s codes. M-A released the participants ID code after data analysis was performed. Due to the nature of the interventions, participantś blinding was not possible.

### Experimental setup and tasks

Each experimental session included completing questionnaires, ultrasonography of the AT and high-density surface electromyography (HD-sEMG) of the triceps surae muscle during plantarflexion isometric contractions at 10%, 40%, and 70% MVC. Details of the experimental session can be found in a previous study from our group ^14^.

For the individuals with NIAT, information regarding their symptoms (NRS and duration of symptoms) was obtained. Pain intensity was recorded on each experimental session. After this assessment, a battery of questionnaires, which included the VISA-A ^40^, Foot and Ankle Ability Measure (FAAM) ^44^, Tampa Scale for Kinesiophobia (TSK) ^45^, Pain Catastrophising Scale (PCS) ^46^, and the International Physical Activity Questionnaire short form (IPAQ-SF) ^47^ was completed. Then, participants laid prone on a Biodex System 3 dynamometer (Biodex Medical System), with their knees extended, the pelvis stabilized with a strap, and the assessed foot tightly strapped on the footplate. The ankle was positioned in 0° of plantarflexion and the axis of the dynamometer was aligned with the inferior tip of the lateral malleolus. In this position, tendon morphomechanical properties were assessed using ultrasonography. HDsEMG electrode grids were placed on the MG, LG and SO muscles in the same position across participants. Then, tendon mechanical parameters were estimated using shear-wave elastography during rest conditions.

Participants underwent a warm-up protocol comprising 3 submaximal isometric plantarflexion contractions during 5 seconds at their perceived 30% of maximal voluntary force. After this warm-up, the maximal voluntary contraction (MVC) was determined during 3 maximal plantarflexion contractions during 5 seconds with 2 minutes of rest between contractions. The highest MVC value obtained was used as the reference maximal torque. Then, the electromyographic activity of the MG, LG, and SO muscles was assessed during two isometric plantarflexion contractions at 10, 40, and 70% MVC (10% MVC/s ramp-up, 10 s hold, 10% MVC/s ramp-down and 30 s rest).

### Ultrasonography

All ultrasound images were acquired using an ultrasonography device equipped with SWE (LOGIQS8, GE Healthcare, Milwaukee, USA). Morphological properties included tendon thickness and cross-sectional area (CSA). Mechanical properties encompassed estimated stiffness. Tendon thickness and CSA were recorded in B-mode with a 16-linear array probe (50 mm, 4-15 MHz), and stiffness was estimated via SWE with a 9-linear array probe (44 mm, 2-8 MHz). An intra-rater/inter-session reliability assessment was performed in six asymptomatic individuals (see details in ^48^) to check for consistency of the ultrasound measures, showing excellent reliability for thickness (ICC:0.99 and 0.99), good reliability for stiffness (ICC:0.90) and moderate reliability for CSA (ICC:0.64).

Morphological tendon properties were determined using an adaptation of the protocol developed by Arya and Kulig ^19^. Tendon thickness and CSA were assessed at 2, 4, and 6 cm from the insertion, in longitudinal and transverse planes respectively.

Details for HD-sEMG electrode grid placement have been reported previously ^14^, consistency of grid placement was made via skin marking with a surgical pen, photographs and the use of landmarks reported in our previous studies ^14, 48^. For AT stiffness assessment, the ankle was set at 0° plantarflexion and the probe positioned longitudinally over the posterior calcaneus, aligned with a mark 4 cm above the AT insertion. A probe holder minimized pressure, and four SWE images were acquired over 12 seconds (one every 2.4 seconds).

### HD-sEMG and torque recordings

The skin was shaved, gently abraded (Nuprep, Skin Prep Gel, Aurora, Colorado) and cleaned with water and dried with paper for HDsEMG electrode preparation. HD-sEMG signals from the MG, LG, and SO muscles were recorded using three two-dimensional (2D) adhesive grids (OT Bioelettronica, Italy) of 13 x 5 equally spaced electrodes (1 mm diameter, inter-electrode distance of 8 mm) filled with conductive paste (AC-CREAM, SPES Medica, Genova, Italy).

HD-sEMG signals were digitized by a 16-bit analogue-digital converter (Quattrocento-OT Bioelecttronica, Torino, Italy), amplified by a factor of 150, sampled at 2048 Hz, and filtered with a band-pass filter (bandwidth: 10-500 Hz, first order, -3 dB) ^49^. HD-sEMG signals were collected in monopolar mode with ground electrodes (WhiteSensor WS, Ambu A/S, Ballerup, Denmark) positioned the head of the fibula and with a wet strap on the thigh of the assessed leg. The torque exerted by the participants was obtained with a Biodex System 3 dynamometer synchronized with the HD-sEMG signals.

### Image analysis

Tendon thickness. Tendon thickness was assessed at 2, 4, and 6 cm from the insertion using ImageJ (http://imagej.nih.gov/ij), averaging three ultrasound images per site. Tendon CSA was measured at the same locations using ultrasound device tools, also averaging three images per site. Tendon stiffness was evaluated using four SWE color maps (2.5 × 1 cm). A 3 mm region of interest was placed along a central reference line in the tendon’s midsection, and stiffness was calculated by averaging values from four consecutive images ^14^.

### HD-sEMG signal analysis

*Motor unit analysis.* The HD-sEMG signals were decomposed into motor unit spike trains with an algorithm based on blind source separation ^28^. The Silhouette metric was used to determine the decomposition accuracy of identified motor units and was set to ≥ 0.90 ^50^. Motor unit properties were assessed by analyzing the whole pool of identified units and further confirmed by a motor unit longitudinal tracking approach ^51^. Motor units were tracked in pairwise fashion between session 1 vs session 2 and session 1 vs session 3 ^51^. A spatial 2D cross-correlation coefficient of >0.75 was used to assume that the motor unit was the same.

In this study, motor unit analysis focused on the effects of CON and ECC exercises on the variables that were found to be altered by NIAT in our previous case–control study^14^. Therefore, we focused on the assessment of mean discharge rate, recruitment and de-recruitment thresholds, and motor unit firing torque relationships. Mean Discharge rate was calculated from the steady phase of the contraction (10s duration). Recruitment and de-recruitment thresholds were defined as the torque (%MVC, or Nm in the case of tracked motor units) when motor units began or ceased firing potentials, respectively ^52^. Missing pulses producing non-physiological firing rates were manually removed and iteratively deleted, and the pulse train was re-calculated ^28^. Motor unit firing rate properties were recorded, analyzed, and reported according to the consensus for experimental design in electromyography: single motor unit matrix ^53^.

### Cross-correlation coefficient and neuromechanical delay

The interactions between neural drive and force generation were examined using cross-correlation analysis. Motor unit discharge times obtained from the decomposition, were summed to generate a cumulative spike train (CST) that depicts cumulative motor unit activity ^28^. Then, a low-pass filter (4^th^ order zero-phase Butterworth, 2 Hz) was applied to the CST and torque signals, followed by high-pass filtering (4^th^ order zero-phase Butterworth, 0.75 Hz) as reported previously ^54^. CST and torque values were cross-correlated to determine similarities in their fluctuations (cross-correlation coefficient) ^28^. The lag obtained from cross-correlation was used to determine the delay between CST and torque (neuromechanical delay).

### Training sessions

Both ECC and CON groups came to our laboratory to perform the training sessions with IC-H, using a Biodex System 3 dynamometer. Participants in the ECC group were asked to perform a warm-up protocol consisting of 3 eccentric plantarflexion contractions at 25% MVC followed by the training protocol which consisted of 4×15 eccentric plantarflexion contractions at 50% MVC (determined at 0° of plantarflexion) with a range of motion from 0° to 30° of plantarflexion. The angular speed was kept at 3°/s (equating to a time-under-tension of 10 seconds) and three minutes of rest were provided between series. Participants in the CON group were asked to complete the same protocol, but with concentric contractions. Visual feedback of the exerted torque was provided in all training sessions and participants were instructed to match the torque output as closely as possible to the target torque during the duration of the contraction. During ECC, the device passively returned the foot to plantarflexion, while during CON, it passively returned the foot to dorsiflexion. The MVC was adjusted for the ECC and CON groups every two weeks of training and was based on the maximal isometric MVC torque produced.

### Follow-up

Participants were contacted via email after 3 and 6 months following the completion of CON and ECC to report NRS and VISA-A scores.

### Statistical analysis

Descriptive statistics were reported as means and SDs, unless otherwise specified. The Shapiro-Wilk test was used to determine the normality of the data, and the Levene test was used to assess the assumption of homogeneity of variance. As these assumptions were met, parametric statistical tests were deemed appropriate. All motor unit analyses were performed using R software (version 4.4.3; R Development Core Team, 2023) by using linear mixed-effects models (LMMs) through the lme4 package (version 1.1.36) ^55^. Data was analysed with LMMs for both the full population of identified motor units as well as for units that were tracked across the interventions. Separate models were created for each muscle to analyse the data independently. The model for mean discharge rate was specified as:

#### Mean Discharge Rate ∼ Exercise x Torque level x Session + (1 | Participant)

This model is interpreted as “the variable of interest (on the left) is predicted by each of the factors on the right. In this case, the variable of interest (Mean Discharge Rate) is the dependent variable, while the independent (explanatory) variables, or “fixed effects,” are exercise group (CON, ECC), force level representing contraction intensity (10%, 40%, and 70% MVIC), and session (session 1, session 2, and session 3). The term (1 | Participant) represents the “random effect,” which accounts for variability due to individual differences. This modeling approach was applied to all outcome variables except for comparisons of demographic data and questionnaires between groups, for which two-way repeated measures ANOVAs were used (factors: group and session). Following model fitting, residual normality was evaluated using the Shapiro-Wilk test. When normality assumptions were violated, outliers in the residuals were identified and excluded based on Cook’s distance with a criterion of four standard deviations ^56^. Fixed effects significance was tested using Type III Wald F tests employing Kenward-Roger degrees of freedom through the ANOVA function in R’s car package (version 3.1.3). Pairwise post-hoc analyses were conducted using Tukey-adjusted comparisons and least-squares contrasts via the emmeans package (version 1.10.7), with LMM outcomes presented as estimated means (M) and 95% confidence intervals (CI). The alpha level for statistical significance was established at p<0.05.

## RESULTS

### Baseline participant characteristics

From the 26 participants, 24 completed the study (12 per group). Participants baseline anthropometrics, pain, questionnaire scores and AT morphomechanical features for the ECC and CON groups are reported in **Table 1**. Differences were only detected in tendon stiffness, with the CON group showing higher stiffness values than ECC.

**Table 1.** Participant baseline characteristics.

|  | ECC group (N=12) | CON group<br>(N=12) | p-value | 95% CI |
| --- | --- | --- | --- | --- |
| Age (years) | 28.83 ± 6.85 | 29.67 ± 10.20 | 0.816 | [-8.19, 6.52] |
| Sex (males/females) | 7/5 | 6/6 | -- | -- |
| Height (cm) | 174.38 ± 8.28 | 171.31 ± 10.62 | 0.437 | [-4.99, 11.14] |
| Weight (kg) | 74.57 ± 19.40 | 74.57 ± 16.54 | 1.00 | [-15.26, 15.26] |
| Laterality (Right/Left) | 11/1 | 11/1 | -- | -- |
| Assessed leg (Right/Left) | 6/6 | 8/4 | -- | -- |
| Bilateral symptoms | 4 | 7 | -- | -- |
| Symptoms duration (months) | 33.42 ± 34.35 | 49.50 ± 40.47 | 0.305 | [-47.86, 15.70] |
| NRS | 2.92 ± 1.16 | 3.25 ± 1.29 | 0.513 | [-1.37, 0.71] |
| VISA-A | 68.67 ± 11.96 | 63.00 ± 19.07 | 0.393 | [-7.81, 19.14] |
| IPAQ-SF | 128.33 ± 71.55 | 121.67 ± 46.73 | 0.789 | [-44.49, 57.83] |
| FAAM (Subscale Activities of Daily Living) | 89.58 ± 7.24 | 81.67 ± 12.18 | 0.066 | [-0.56, 16.40] |
| FAAM (Subscale Sports) | 70.42 ± 15.66 | 65.75 ± 18.36 | 0.510 | [-9.78, 19.11] |
| PCS | 11.75 ± 9.23 | 13.42 ± 12.81 | 0.718 | [-11.12, 7.78] |
| TSK | 37.08 ± 4.42 | 37.17 ± 9.04 | 0.977 | [-6.11, 5.94] |
| Length (cm) | 21.16 ± 4.01 | 21.19 ± 2.20 | 0.980 | [-2.77, 2.71] |
| Thickness 2 cm (cm) | 0.34 ± 0.05 | 0.37 ± 0.07 | 0.396 | [-0.07, 0.03] |
| Thickness 4 cm (cm) | 0.42 ± 0.09 | 0.49 ± 0.11 | 0.103 | [-0.16, 0.02] |
| Thickness 6 cm (cm) | 0.43 ± 0.05 | 0.51 ± 0.16 | 0.134 | [-0.17, 0.02] |
| CSA 2 cm (cm<sup>2</sup>) | 0.43 ± 0.07 | 0.49 ± 0.08 | 0.078 | [-0.12, 0.01] |
| CSA 4 cm (cm<sup>2</sup>) | 0.41 ± 0.07 | 0.47 ± 0.10 | 0.102 | [-0.13, 0.01] |
| CSA 6 cm (cm<sup>2</sup>) | 0.46 ± 0.07 | 0.50 ± 0.10 | 0.266 | [-0.11, 0.03] |
| Stiffness (kPa) | 63.58 ± 6.45 | 72 ± 90 ± 6.3 | 0.002* | [-14.71, -3.92] |

Values are presented as mean ± SD (Standard deviation). 95% Confidence intervals are reported. ECC, eccentric; CON, concentric; NRS, Numerical Rating Scale; N/A, non-applicable. \* Indicates significant statistical difference, p<0.05.

### Pain, questionnaire and ultrasonographic results

NRS pain and VISA-A scores between ECC and CON groups at baseline (week 1), weeks three, six, 3-month follow-up and 6-month follow-up are presented in **Figure 1**. NRS scores between ECC and CON decreased similarly in both groups with time (Time effect: F= 47.61, P<0.0001). VISA-A scores increased similarly between groups (Time effect: F=23.18, P<0.001), with a significant increase in scores from week 6.

**Figure 1.**
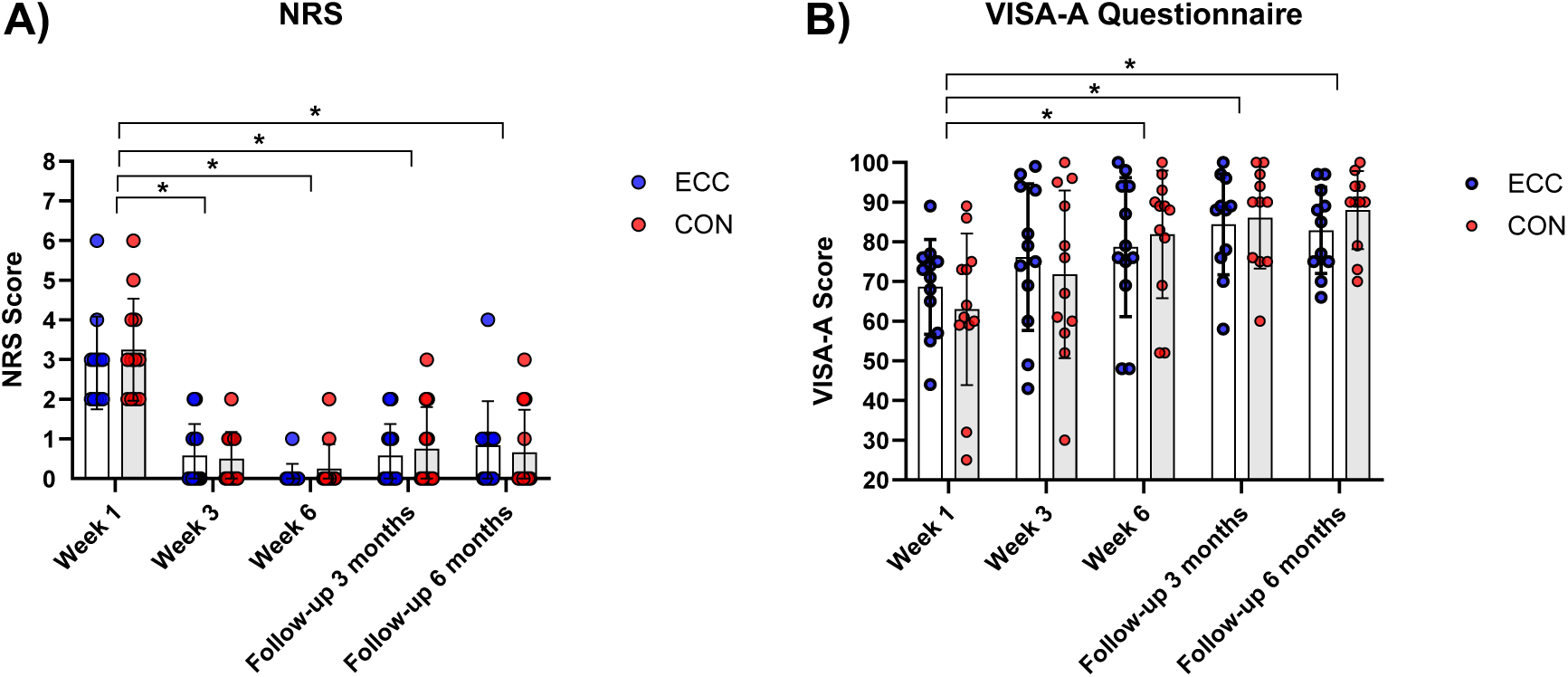
A) Numerical Rating Scale (NRS) scores between eccentric (ECC) and concentric (CON) groups at week 1, week 3, week 6, follow-up 3 months and follow-up 6 months. B) Victorian Institute of Sports Assessment – Achilles (VISA-A) questionnaire scores between eccentric (ECC) and concentric (CON) groups at week 1, week 3, week 6, follow-up 3 months and follow-up 6 months. * Statistical differences between assessment times, p<0.05.

Other questionnaire scores are presented in **Table 2**. IPAQ scores showed no significant changes across the intervention (P=0.27). FAAM scores demonstrated a significant increase over time (Time effect: F=7.24, P=0.0038), with no differences between groups (Group effect: P=0.80). Similarly, PCS scores decreased over time (Time effect: F=6.38, P=0.01), but no group differences were found (Group effect: P = 0.87). TSK scores also decreased over time (Time effect: F=9.66, P=0.002), with no significant differences between groups (Group effect: P=0.70).

**Table 2.** IPAQ, FAAM, PCS, TSK questionnaire scores and ultrasound imaging results for concentric and eccentric groups.

|  | Baseline Con | 3wk Con | 6wk Con | Baseline ECC | 3wk ECC | 6wk ECC |
| --- | --- | --- | --- | --- | --- | --- |
| Questionnaire results |  |  |  |  |  |  |
| IPAQ-SF | 121.67 (44.74) | 132.08 (72.38) | 155 (80.49) | 128.33 (68.5) | 127.92 (67.8) | 131.25 (69.07) |
| FAAM (ADL) | 81.67 (11.65) | **85 (12.23)** | **91.08 (11.08)** | 89.58 (6.93) | **91.08 (5.48)** | **92.42 (6.7)** |
| FAAM (SPORTS) | 65.75 (17.57) | **72 (17.08)** | **80.58 (14.24)** | 70.42 (15.57) | **74.08 (16.12)** | **77.75 (17.91)** |
| PCS | 13.42 (12.26) | **8.33 (8.19)** | **9.17 (8.83)** | 11.75 (8.83) | **8.75 (8.96)** | **8.58 (9.27)** |
| TSK | 37.2 (8.7) | **32.92 (9.7)** | **32 (9.2)** | 37.08 (4.23) | **35.17 (7.1)** | **33.17 (7.4)** |
| Maximal torque results |  |  |  |  |  |  |
| MVC torque (Nm/Kg) | 1.13 (0.35) | **1.22 (0.40)** | **1.37 (0.45)** | 1.18 (0.26) | **1.30 (0.25)** | **1.45 (0.26)** |
| Ultrasound imaging results |  |  |  |  |  |  |
| Thickness 2cm (cm) | 0.49 (0.07) | 0.48 (0.07) | 0.48 (0.1) | 0.44 (0.07) | 0.44 (0.07) | 0.43 (0.07) |
| Thickness 4cm (cm) | 0.475 (0.1) | 0.47 (0.09) | 0.47 (0.1) | 0.41 (0.06) | 0.42 (0.07) | 0.42 (0.06) |
| Thickness 6cm (cm) | 0.49 (0.1) | 0.49 (0.1) | 0.49 (0.1) | 0.46 (0.07) | 0.47 (0.07) | 0.46 (0.06) |
| CSA - 2cm (cm<sup>2</sup>) | 0.37 (0.07) | 0.36 (0.06) | 0.37 (0.06) | 0.35 (0.05) | 0.34 (0.04) | 0.34 (0.05) |
| CSA - 4cm (cm<sup>2</sup>) | 0.49 (0.10) | 0.48 (0.1) | 0.48 (0.1) | 0.42 (0.09) | 0.42 (0.08) | 0.42 (0.08) |
| CSA - 6cm (cm<sup>2</sup>) | 0.51 (0.15) | 0.5 (0.14) | 0.50 (0.14) | 0.43 (0.05) | 0.43 (0.05) | 0.43 (0.05) |
| Stiffness (Kpa) | **72.89 (6.0) #** | **76.59 (5.6) #** | **82.67 (4.71)** | **63.58 (6.2) #** | **66.89 (6.53) #** | **79.25 (4.6)** |
| Normalised Stiffness (%) | ----- | **5.08 (8.76)** | **13.41 (9.05) \*** | ----- | **5.20 (6.67)** | **24.64 (6.93) \*** |

Questionnaire and ultrasound imaging results across the intervention (baseline, week 3 and week 6) for the concentric (CON) and eccentric (ECC) training groups. Significant differences between week 3 versus baseline, and week 6 versus baseline are highlighted in bold (p<0.05). # Denotes a significant difference between CON and ECC at baseline and week 3 (p<0.05). \* Denotes a significant difference between CON and ECC at week 6 (p=0.01).

Ultrasound imaging results can be also seen in **Table 2**. Overall, thickness and cross-sectional area were not changed by the interventions (P>0.18 in all cases). Due to the differences in stiffness at baseline between ECC and CON groups, the percental stiffness change at weeks 3 and 6 relative to baseline scores was calculated. Overall, stiffness increased differently across groups and times (Interaction effect: F=7.01, P=0.02) with higher stiffness in the ECC compared to the CON group at week 6 (P=0.01, 95% CI=2.66% to 19.09%).

### Strength

A significant session effect was observed for MVC torque (F=17.53, p<0.0001) with no differences between groups (p=0.89), post hoc tests revealed significant increases from baseline to session 2 (MD= - 8.15 Nm, 95% CI [-15.90, -0.36]), from baseline to session 3 (MD= -18.95 Nm, 95% CI [-26.70, -11.17]), and from session 2 to session 3 (MD= -10.81 Nm, 95% CI [-18.60, -3.02]).

### Motor unit identification and tracking

Across the intervention and considering all force levels, an average of 21 (4), 10 (1), and 17 (3) motor units were identified per participant in the ECC group for the MG, GL, and SOL muscles, respectively. In the CON group, the averages were 18 (2), 10 (1), and 17 (1) motor units for the same muscles. There were no significant differences in the total number of identified motor units between groups (p=0.4). Regarding motor unit tracking across all force levels, in the ECC group, an average of 6 (3), 2 (1), and 2 (1) motor units were successfully tracked between sessions 1 and 2 for MG, GL, and SOL, respectively. Between sessions 1 and 3, the averages were 5 (2), 3 (1), and 3 (1) for the same muscles. In the CON group, 4 (1), 2 (1), and 3 (0) motor units were tracked between sessions 1 and 2, and 5 (2), 2 (1), and 2 (1) were tracked between sessions 1 and 3 for MG, GL, and SOL, respectively. The average spatial cross-correlation coefficient of all tracked motor units in both groups was 0.80 (0.06) between sessions 1 and 2, and 0.78 (0.05) between sessions 1 and 3.

### Motor unit firing results: recruitment and de-recruitment thresholds

Motor unit recruitment and de-recruitment thresholds demonstrated muscle-and exercise-specific adaptations, as shown in **Figure 2**. In the MG muscle, a significant exercise × session interaction was observed for recruitment thresholds (F=6.24, p=0.002), with an increase in the CON group from session 1 to 3 (MD= -1.72, 95% CI [-3.24, -0.20]) and in the ECC group from session 2 to 3 (MD= -1.83, 95% CI [-3.24, -0.42]). These changes were consistent with tracked motor units, which showed a significant increase in recruitment thresholds in both groups between sessions 1 and 3 (p<0.0001, MD= -5.05 Nm, 95% CI [-6.50, -3.60]). For MG de-recruitment thresholds, a significant exercise × force × session interaction emerged (F=3.05, p=0.016), with increases in the CON group at high loads (70% MVC) between sessions 1 and 3 (MD= -4.82, 95% CI [-8.63, -1.01]) and sessions 2 and 3 (MD= -6.57, 95% CI [-10.76, -2.38]), paralleled by increases in tracked units across both groups from session 1 to 3 (p=0.003, MD= -2.10 Nm, 95% CI [-3.46, -0.73]). In GL, a significant exercise × session interaction was also found for recruitment thresholds (F=3.57, p=0.028), with the CON group showing increases from session 1 to 2 (MD= -4.17, 95% CI [-6.12, - 2.23]) and from session 1 to 3 (MD= -5.09, 95% CI [-7.07, -3.10]), whereas the ECC group showed a significant change only from session 1 to 3 (MD= -2.96, 95% CI [-5.05, -0.87]). Tracked motor unit data supported these findings, showing a significant increase in recruitment thresholds in the CON group between sessions 1 and 2 (exercise × session interaction: p=0.04, MD = -5.03 Nm, 95% CI [-9.08, -0.97]), and a comparable increase in both groups from session 1 to 3 (session effect: p < 0.0001, MD = -7.10 Nm, 95% CI [-9.76, -4.43]). GL de-recruitment thresholds exhibited a significant exercise × force level × session interaction (F = 7.54, p < 0.0001), with marked increases in the ECC group at high loads from session 1 to 2 (MD= -10.39, 95% CI [-15.54, -5.24]) and from session 1 to 3 (MD= -13.06, 95% CI [-18.22, -7.90]), this was backed by tracked data, showing increases between sessions 1 and 3 in ECC at 70% MVC (p=0.001, MD= -4.98 Nm, 95% CI [-1.27, -8.68]). In SOL, recruitment thresholds also showed a significant exercise × force level × session interaction (F= 2.85, p= 0.023), with increases in the CON group at 40% MVC from session 1 to 2 (MD= -3.28, 95% CI [-6.10, -0.47]) and from session 1 to 3 (MD= -5.86, 95% CI [-8.72, -3.00]), and at 70% MVC from session 2 to 3 (MD= -3.03, 95% CI [-6.01, -0.06]). Tracked motor unit data showed similar recruitment threshold increases in both groups from session 1 to 3 (p=0.0001, MD= -4.36 Nm, 95% CI [-6.58, -2.14]). Soleus de-recruitment thresholds increased progressively in both groups across sessions (F = 27.25, p < 0.0001), with significant differences between sessions 1 and 2 (MD= -1.88, 95% CI [-2.87, - 0.88]), 1 and 3 (MD= -3.08, 95% CI [-4.06, -2.10]), and 2 and 3 (MD= -1.21, 95% CI [-2.13, -0.29]). These changes were echoed in the tracked motor units, with a significant increase in de-recruitment thresholds between sessions 1 and 3 across both groups (F = 21.32, p < 0.0001, MD = -4.93 Nm, 95% CI [-7.04, -2.83]). Tracked motor unit recruitment and de-recruitment threshold results are provided in **Supplementary Figure 2**.

**Figure 2.**
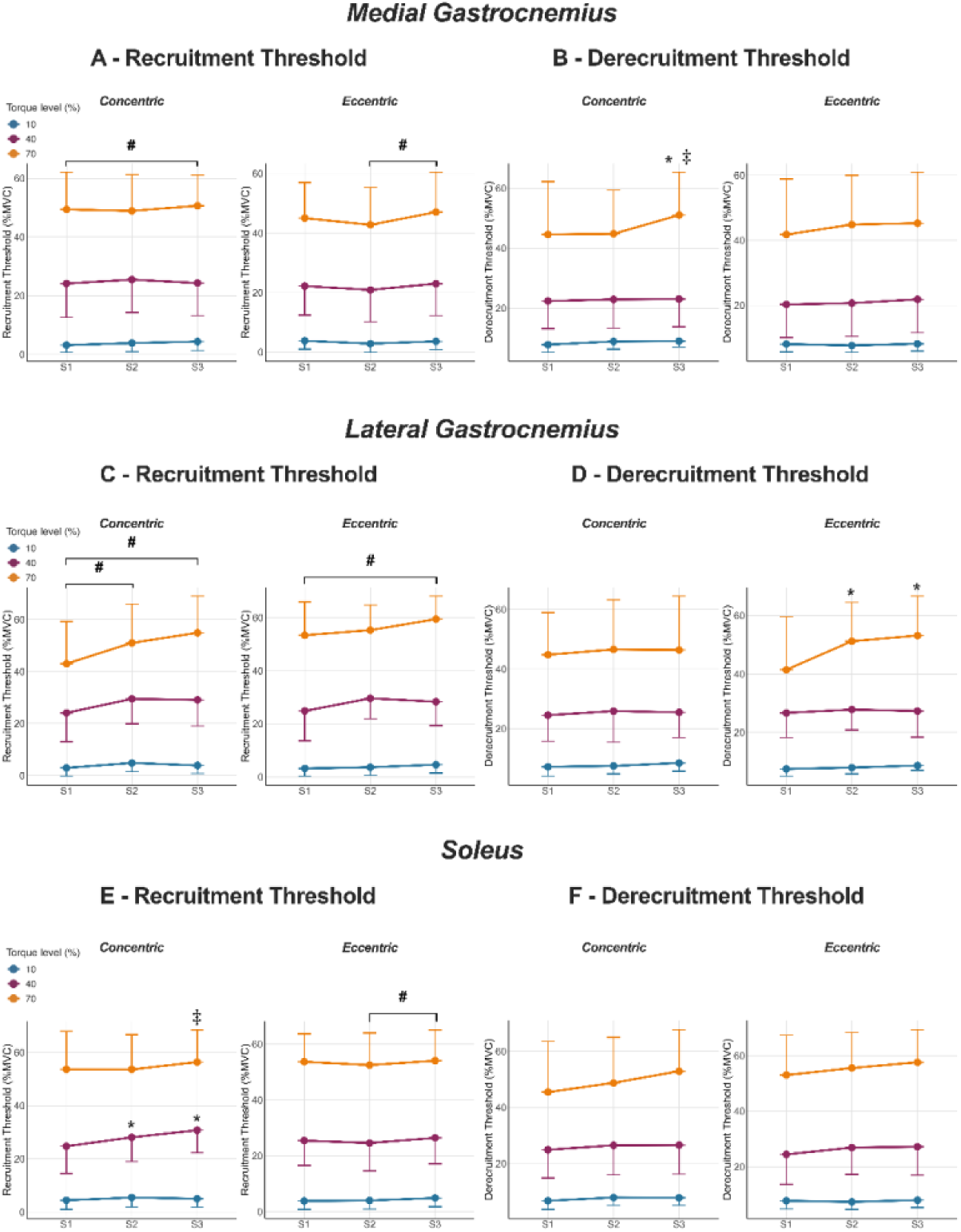
Changes in recruitment and de-recruitment torque thresholds for gastrocnemius medialis, gastrocnemius lateralis and soleus muscles across the three sessions (S1, S2, S3) and three force levels (10%, 40% and 70% MVC) for all identified motor units. A) Recruitment thresholds for gastrocnemius medialis. B) De-recruitment thresholds for gastrocnemius medialis. C) Recruitment thresholds for gastrocnemius lateralis. D) De-recruitment thresholds for gastrocnemius lateralis. E) Recruitment thresholds for Soleus. F) De-recruitment thresholds for soleus. All results are presented as means and standard deviations. #, denotes significant session effects (p<0.05). *, denotes significant differences from baseline (S1), at each independent force level (p<0.05). ‡, Denotes significant differences from S2, at each independent force level (p<0.05).

### Motor unit firing results: Mean Discharge rate

There were muscle and exercise-dependent changes in discharge rate between groups as observed in **Figure 3**. Medial gastrocnemius showed a significant exercise × force × session interaction (F=4.79, p=0.0008). Specifically, there was an increase in MG discharge rate at 40% MVC in the CON group from session 1 to session 3 (MD= -0.72, 95% CI [-1.23, -0.21]) and from session 2 to session 3 (MD= -0.63, 95% CI [-1.12, -0.15]), while the ECC group showed a decrease from session 1 to session 2 at 40% MVC (MD= 0.61, 95% CI [0.13, 1.10]). These results were confirmed from tracked motor units where reductions in discharge rate between sessions 1 and 2 (p<0.0001, MD= 0.97Hz, 95% CI [0.56, 1.37]) and 1 and 3 (p=0.004, MD= 0.54Hz, 95% CI [0.07, 1.01]) were observed for the ECC group, with the CON group showing an increase between sessions 1 and 3 (p<0.001, MD= 0.75, 95% CI [0.4928 to 1.015]. In GL, a significant force × session interaction was observed (F = 7.07, p < 0.0001). Post hoc analysis revealed a dichotomous response pattern across contraction intensities. At moderate loads (40% MVC), discharge rate increased from session 1 to session 3 (MD= -0.56, 95% CI [-1.08, -0.05]). However, during higher load contractions (70% MVC), a different pattern was observed with reduced discharge rate from session 1 to session 2 (MD= 0.73, 95% CI [0.11, 1.34]) and from session 1 to session 3 (MD= 0.77, 95% CI [0.14, 1.39]) in both groups. These findings were confirmed for tracked units as a significant increase in GL discharge rate at 40% and a decrease in discharge rate at 70% MVC was observed between sessions 1 and 3 in both groups (session x force interaction, F=5.55, p=0.043). Finally, for SO, a significant exercise × force level × session interaction was observed (F=2.97, p=0.018), with increases in discharge following ECC at moderate loads (40% MVC) from session 1 to session 3 (MD= -1.14, 95% CI [-2.04, -0.23]) and at higher loads (70% MVC) from sessions 1 to 2 (MD= -1.11, 95% CI [-2.21, -0.02]). This finding was partially supported by tracked motor unit data, which revealed a significant main effect of session. Specifically, discharge rates were significantly higher in session 2 (F=4.78, p=0.03, MD= -0.55 Hz, 95% CI [-1.05, -0.06]) and session 3 (F=5.61, p=0.019, MD= -0.64 Hz, 95% CI [-1.17, -0.11]) in both groups. Tracked motor unit discharge rate results can be found in **Supplementary figure 3**.

**Figure 3.**
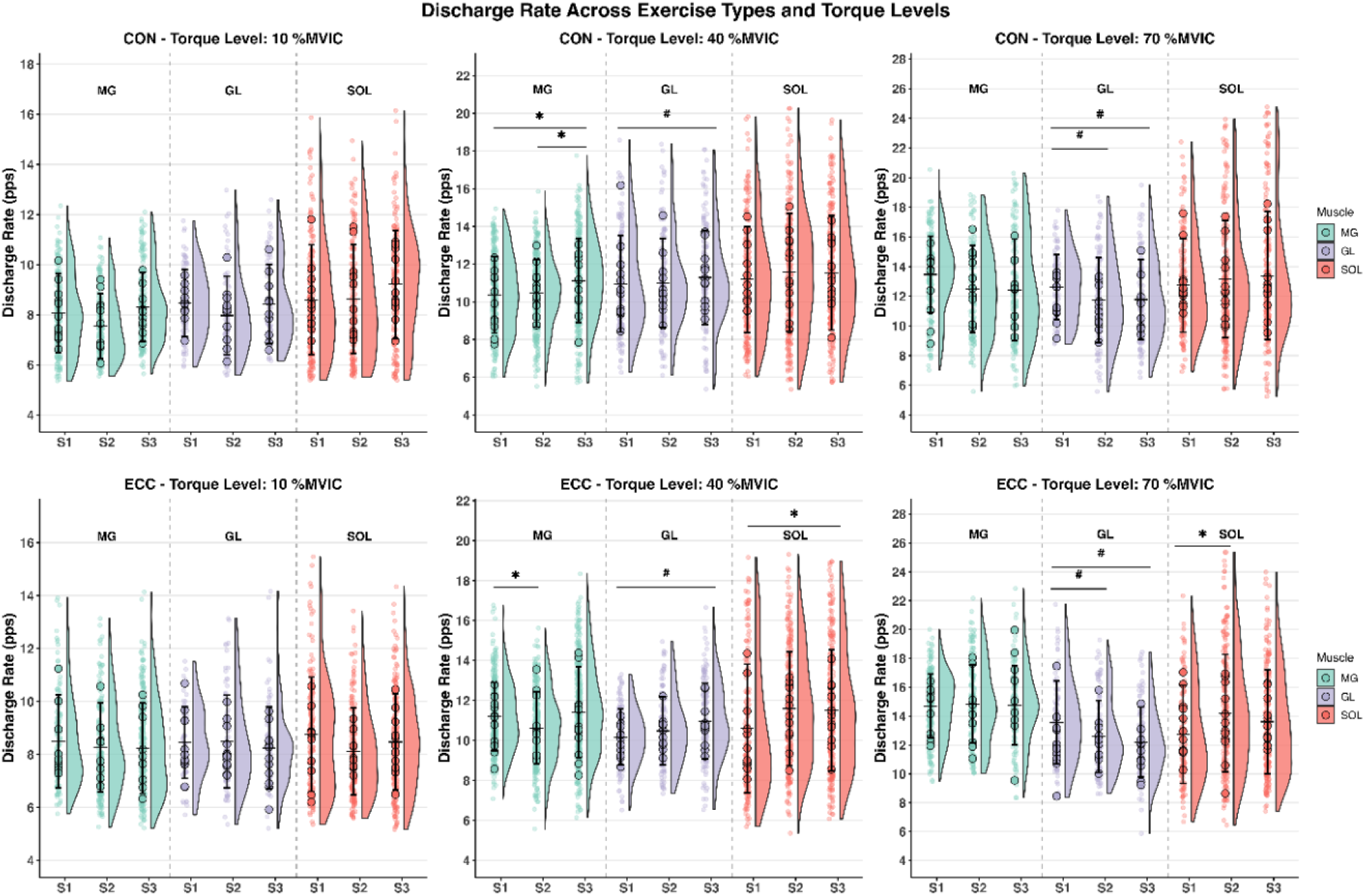
Changes in mean discharge rate for for the full population of identified motor units in gastrocnemius medialis (MG), gastrocnemius lateralis (GL) and soleus (SOL) muscles across the three sessions (S1, S2, S3) and three force levels (10%, 40% and 70% MVC) for concentric (CON) and eccentric (ECC) exercises. CON exercise results are reported in the top panels: 10% MVC, left, 40% MVC, centre and 70% MVC, right. ECC exercise results are reported in the bottom panels: 10% MVC, left, 40% MVC, centre and 70% MVC, right. #, denotes significant session effects (p<0.05). *, denotes significant differences for pairwise comparisons for exercise x session interactions (p<0.05).

### Torque-firing rate relationships and neuromechanical delay

There was a significant reduction in GL cross-correlation (CST versus torque) across both interventions (session effect: F=3.77, p=0.025) with a significant reduction in CST-torque cross-correlation from session 1 to session 3 (MD= 0.04, 95% CI [0.01, 0.08]). Furthermore, there was a significant main effect of session for SO’s muscle neuromechanical delay (F=3.53, p=0.031) with a significant decrease in neuromechanical delay from sessions 1 to 3 (MD= 46.3 ms, 95% CI [1.24, 91.4]). Torque-firing rate relationship results can be found in **Figure 4**.

**Figure 4.**
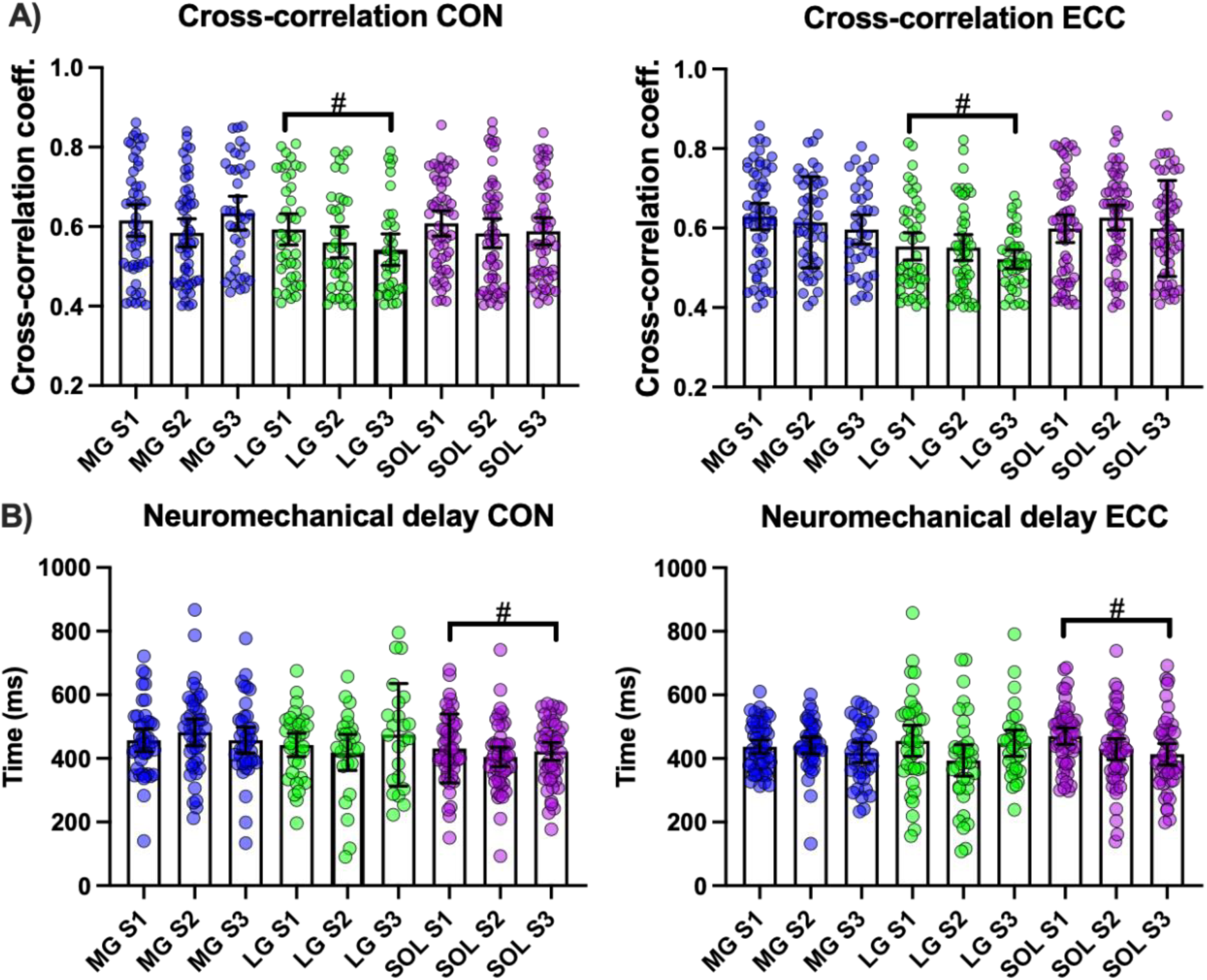
Changes in motor unit firing–torque relationships across the three sessions (S1, S2, S3) during concentric (CON) and eccentric (ECC) exercises in the Medial Gastrocnemius (MG), Lateral Gastrocnemius (LG), and Soleus (SOL) muscles. (A) Cumulative spike train–torque cross-correlation for CON (left) and ECC (right). (B) Neuromechanical delay for CON (left) and ECC (right). #: Indicates a significant effect of session (p < 0.05).

## DISCUSSION

This is the first study to demonstrate that a 6-week torque-feedback training intervention, based on either controlled CON or ECC exercise, leads to significant improvements in pain and function, regardless of the protocol used. However, these symptomatic improvements were accompanied by distinct neuromechanical adaptations between training modalities, including load-and muscle-specific changes in motor unit firing activity and differential alterations in muscle stiffness as estimated by SWE. Collectively, these findings provide novel insights into the neuromechanical mechanisms underlying symptom relief and functional enhancement following CON and ECC training in individuals with NIAT.

### Differences between training protocols

Although eccentric exercise has been widely used in the management of NIAT, the mechanisms underlying its effectiveness in improving tendon-related symptoms remain unclear ^57^. Exercise protocols employing eccentric or concentric contractions—or a combination of both—have been successfully applied in individuals with NIAT ^8, 26^. This has led to increased scrutiny of the idea that the tendon’s response to mechanical loading depends on the type of muscle contraction performed ^32, 35^. To investigate this further, we designed our intervention to isolate eccentric and concentric contractions using an isokinetic dynamometer. Using this approach, we found that both CON and ECC were effective in increasing muscle strength, reducing pain, and improving patient-reported outcomes. It is possible that factors unrelated to contraction type but specific to our intervention protocol (i.e., prolonged time-under-tension, visual feedback for torque control and individualized load progression) contributed to the early improvement in symptoms. These findings support the idea that controlled tendon loading, rather than the specific type of muscle contraction, plays a key role in the effective management of NIAT. A recent review reported that eccentric exercise was the most effective intervention for pain reduction in individuals with NIAT^58^. However, the number of studies was limited, and the overall risk of bias was high. Moreover, most of these studies did not isolate eccentric and concentric contractions. Comparisons often involved single-leg heel-drop exercises (eccentric) versus heel-raise exercises (concentric), where it is difficult to control for gravitational effects and isolate contraction types accurately. This makes it challenging to determine whether the observed benefits were due to eccentric loading alone or a combination of contraction types. Our findings suggest that standardization of contraction mode is crucial for making meaningful comparisons between training interventions. Supporting this, Franchi et al. ^59^ used a similar protocol and found that quadriceps strength improved similarly following both concentric and eccentric training over an 8-week period. This was unexpected given that most previous studies report greater gains in maximal strength following eccentric training. This discrepancy may highlight the importance of isolating contraction types and standardizing mechanical load when evaluating training outcomes. Nevertheless, it is important to consider that such similar outcomes may only be observed following short-term interventions. Given the differences we observed in neuromuscular and tendon adaptations between contraction types, it is plausible that concentric and eccentric interventions may lead to distinct clinical outcomes over time. However, longer-term interventions are needed to confirm this hypothesis.

### Morpho-mechanical properties of the AT following exercise

Eccentric exercise may improve pain and function in individuals with NIAT by enhancing tendon structure, including increased collagen synthesis and fiber realignment, which can boost tendon strength and stiffness ^60^. However, it remains unclear why concentric contractions do not produce similar adaptations, given that Achilles tendon elongation and stiffness during concentric heel-raise and drop exercises seem to be comparable to those in eccentric contractions ^61^. Our results showed no significant changes in Achilles tendon morphology in either group over time. These findings align with current evidence suggesting that morphological adaptations of the tendon in response to loading typically require longer interventions (i.e., >12 weeks) ^62^. Still, findings in individuals with NIAT remain mixed, with some studies reporting reductions in tendon thickness and CSA, while others observe no change following similar interventions ^62^. In contrast, our data showed increased AT stiffness over time, with ECC producing greater gains after six weeks, suggesting that short-term changes likely reflect material property adaptations rather than morphological changes. The greater stiffness observed with ECC may be due to higher force production during lengthening contractions, which places more stress on passive structures like tendons and connective tissue than shortening contractions ^63^.

### Motor unit firing rate properties between ECC and CON groups

Our findings revealed that motor unit firing properties varied depending on the exercise modality. Both training protocols led to an overall increase in motor unit recruitment thresholds. However, the responses in de-recruitment thresholds differed, particularly at higher force levels. Specifically, MG motor units in the CON group were de-recruited at higher forces, while LG motor units in the ECC group also showed de-recruitment at higher forces. Additionally, we observed opposite adaptations in the mean discharge rate of the MG: it decreased in the ECC group but increased in the CON group. In contrast, both groups showed a rise in the discharge rate of SO and a reduction in the discharge rate of the GL at high force levels. These neuromuscular adaptations may underlie the improvements in pain and function observed in both intervention groups. In our recent case-control study, we reported that individuals with NIAT exhibited increased firing rates and reduced de-recruitment thresholds of the GL at high force levels (70%), suggesting dysfunctional motor unit behavior^14^. Notably, both interventions in the present study reduced GL discharge rate at high forces, but only the ECC protocol also increased the de-recruitment threshold of this muscle. These findings may have important implications for plantarflexion force generation. Over-reliance on the GL at high forces, which is the smallest of the plantarflexor muscles—may be maladaptive in NIAT. The interventions may have facilitated a redistribution of load toward the larger plantarflexor muscles, specifically SO and MG. This reorganization of muscle activity may have been driven, in part, by the reductions in pain observed following the training interventions. Pain is known to alter the direction of joint force vectors ^64^, which can lead to imbalanced tendon loading, as commonly seen in individuals with NIAT. By alleviating pain, the interventions may have contributed to more optimal muscle recruitment patterns, improving both the efficiency of neural drive transmission and force transfer to the tendon. Importantly, the increase in LG’s motor unit de-recruitment threshold at high forces following ECC may allow reducing the duration of activity of fast-fatigable motor units in this muscle ^52^, representing another positive neuromuscular change that could enhance fatigue resistance and force efficiency during sustained contractions.

The contrasting adaptations in MG discharge rate—decreasing after ECC but increasing after CON—are particularly noteworthy. Lengthening contractions are known to be metabolically efficient and require less neural drive than shortening contractions ^65^. Eccentric contractions also allow for greater force generation due to enhanced contributions from passive elastic structures like titin and connective tissue, which reduce the need for active cross-bridge cycling ^63^. Consequently, eccentric contractions produce more force per cross-bridge, reducing the number of cross-bridges needed and resulting in lower motor unit discharge rates for a given force output ^66^. We previously reported that motor unit twitch force increases when a muscle is lengthened ^29^. Therefore, the repeated exposure to lengthening contractions in the ECC group likely enhanced motor unit twitch force and consequently reduced the neural drive to the medial gastrocnemius. Furthermore, it is plausible that the greater increase in tendon stiffness observed following ECC training contributed to the reductions in MG motor unit firing rates. We recently reported a direct relationship between tendon stiffness and motor unit discharge rate ^48^. Therefore, the greater stiffness induced by ECC may have enhanced the mechanical efficiency of the muscle–tendon unit, reducing the neural drive required to produce force. However, the fact that this effect was observed only in the MG warrants further consideration. Evidence suggests an uneven distribution of neural drive across the triceps surae^67^, with the MG exhibiting the highest activation during plantarflexion tasks. As a result, it is likely that the MG was more substantially affected by the intervention, which may explain the divergent responses observed between muscles and training protocols.

Seminal studies have shown that eccentric contractions induce recruitment of higher threshold motor units ^68^. Our findings partly align with previous observations, showing increased recruitment thresholds after ECC —but also after CON. This suggests that both protocols may have increased the recruitment of higher-threshold motor units, though this alone doesn’t fully explain the change, as tracked units also showed increased thresholds post-training. While prior studies on isometric training of the tibialis anterior reported reduced recruitment thresholds ^69^—possibly due to increased electromechanical delay and reduced muscle stiffness—our results showed increased tendon stiffness with both protocols, especially ECC. This likely improved force transmission and neural-to-mechanical efficiency, contributing to higher twitch force and recruitment thresholds. However, since recruitment thresholds rose across all muscles, but discharge rate changes between protocols were seen only in the MG, this indicates muscle-specific adaptations within the triceps surae. Despite these differences, both protocols resulted in similar gains in maximal torque. Longer-term studies are needed to determine whether such muscle-specific changes lead to different outcomes in force production over time.

### Motor unit firing versus torque relationships

Motor unit firing-torque relationships offer insight into individual muscle contributions within synergistic groups. In individuals with NIAT, we previously found that LG showed low CST-torque relationships at low forces, with increased contribution at higher forces. Following both interventions, GL CST-torque relationships decreased, suggesting reduced activity—consistent with our mean firing rate findings. Altered force distribution among MG, LG, and SO has been implicated in Achilles tendinopathy ^13, 70, 71^. Crouzier et al. and Contreras-Hernandez et al. ^14^ observed that the contribution of the LG to the total plantarflexion torque is lower in individuals with NIAT compared to controls at low forces ^72^. In our previous study, individuals with NIAT showed increased LG contribution to plantarflexion torque at 70% MVC. The current results suggest a shift in activation toward MG and SOL, likely due to changes in their discharge rates, increasing reliance on muscles with larger cross-sectional areas for force transmission. This is supported by reduced neuromechanical delay in SOL, possibly driven by higher discharge rates and increased tendon stiffness, which may have improved the efficiency of force transmission to the tendon.

### Limitations and future developments

Several methodological considerations should be noted. First, participants in both ECC and CON groups reported low to moderate pain, so findings may not generalize to those with more severe symptoms. Second, blinding was not possible due to the need to explain contraction types during training. Third, tendon stiffness was measured only at rest, as ankle movement during moderate to high isometric contractions prevented reliable assessment. Simultaneous evaluation of tendon stiffness and motor unit firing may offer deeper insight into neuromuscular adaptations. Lastly, future studies should examine whether changes observed during isometric tasks also occur during dynamic plantarflexion, aligning more directly with the training stimulus.

## CONCLUSIONS

This study demonstrates that 6-week torque-feedback training intervention, involving either controlled eccentric or concentric exercises, leads to comparable reductions in pain and improvements in self-reported outcomes in individuals with NIAT. These clinical improvements were accompanied by divergent changes in tendon stiffness and muscle- and load-dependent alterations in motor unit behavior across the triceps surae. Notably, the MG exhibited protocol-specific changes in discharge rates, increasing or decreasing depending on the type of contraction applied. Importantly, both exercise modalities were effective in reversing previously reported maladaptive motor unit firing patterns, such as excessive activation of GL at high force levels and elevated GL CST–torque coupling. These findings support the idea that precise load control, rather than contraction type, is a key factor in the effective management of NIAT. However, further research involving longer intervention periods is needed to determine whether the observed differences in stiffness and motor unit behavior between concentric and eccentric training protocols lead to divergent long-term clinical outcomes.

## DATA AVAILABILITY STATEMENT

The data and analysis codes are available from the corresponding author, EM-V, upon reasonable request.

## ACKNOWLEDGEMENTS

This work was supported by ANID PhD Scholarship awarded by the Government of Chile. Recipient: Ignacio Contreras-Hernandez, Scholarship ID number: 72200295. Francesco Negro was funded by the European Research Council Consolidator Grant INcEPTION (contract no. 101045605). Eduardo

Martinez-Valdes is supported by an Orthopaedic Research UK Early Career Research Fellowship (ORUK ref-574).

## AUTHOR CONTRIBUTIONS

IC-H, DF and EM-V conceived and designed research; IC-H and MA performed experiments; IC-H, MA, DJ-G and EM-V analyzed data; IC-H, MA and EM-V interpreted results of experiments; EM-V and MA prepared figures; IC-H and EM-V drafted manuscript; IC-H, EM-V, DF, DJ-G, FN and MA edited and revised manuscript; IC-H, EM-V, DF, FN, DJ-G and MA approved final version of manuscript.

