## Supplementary figures and images for "Eccentric and Concentric Torque Feedback Training Induce Similar Clinical Improvements but Distinct Triceps Surae Motor Unit Adaptations in Non-Insertional Achilles Tendinopathy: A Randomized Controlled Trial"

### Supplementary figure 1

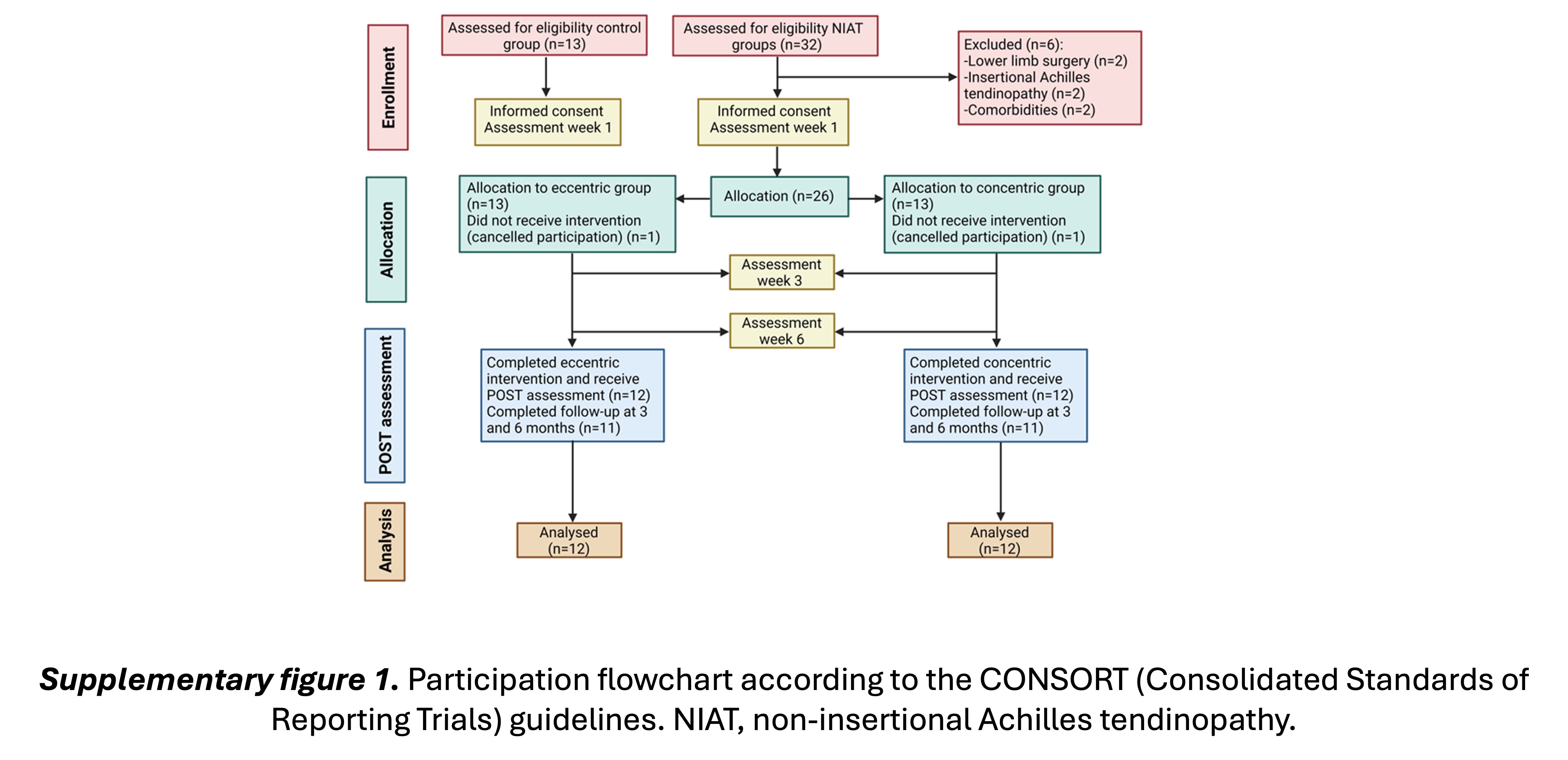
