## Supplementary figure 2 for "Eccentric and Concentric Torque Feedback Training Induce Similar Clinical Improvements but Distinct Triceps Surae Motor Unit Adaptations in Non-Insertional Achilles Tendinopathy: A Randomized Controlled Trial"

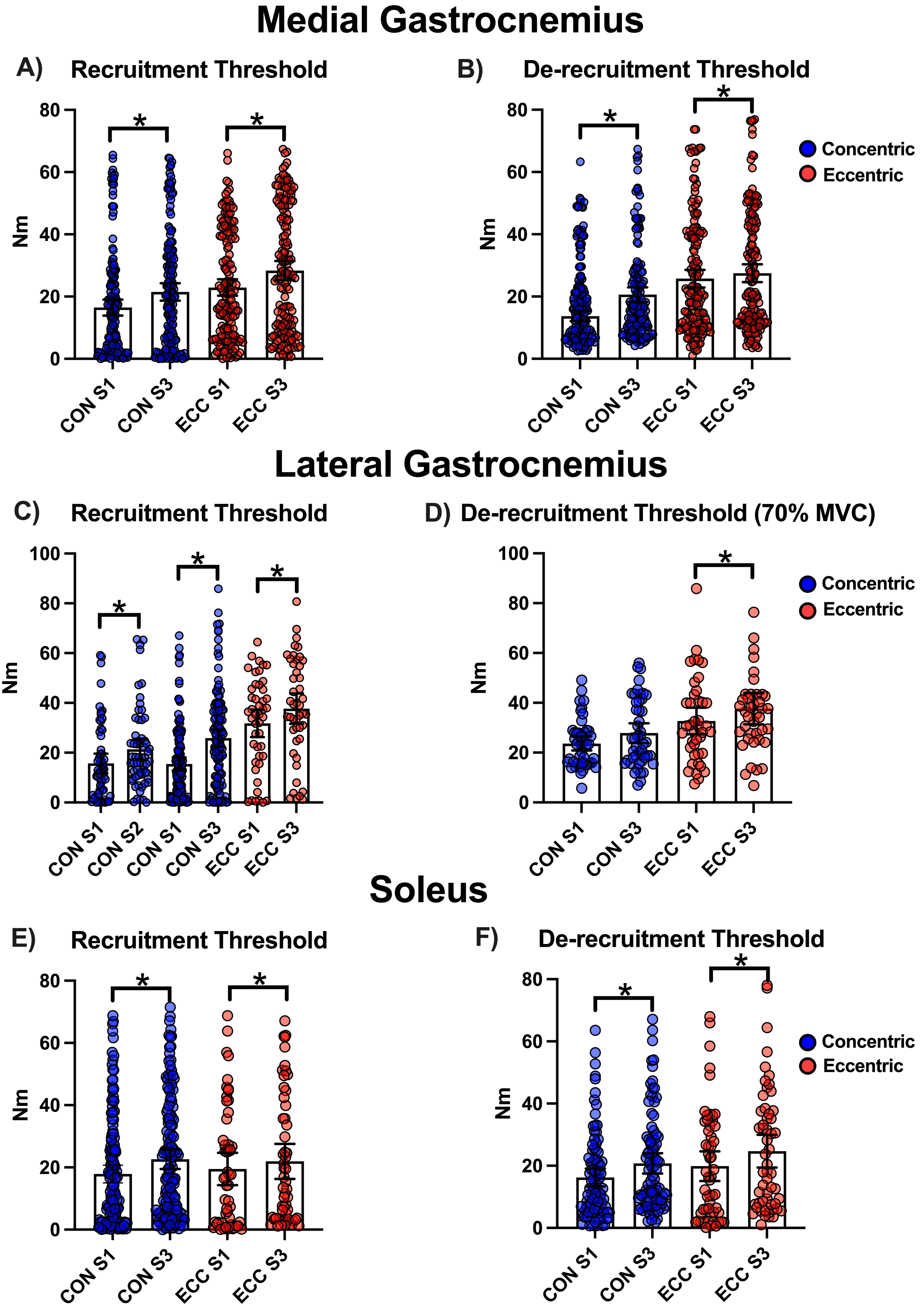


**Supplementary figure 2.** Recruitment and de-recruitment thresholds for tracked motor units in absolute torque values (Nm). A) Recruitment thresholds for gastrocnemius medialis. B) De-recruitment thresholds for gastrocnemius medialis. C) Recruitment thresholds for gastrocnemius lateralis. D) De-recruitment thresholds for gastrocnemius lateralis. E) Recruitment thresholds for Soleus. F) De-recruitment thresholds for soleus. CON, concentric training. ECC, eccentric training. S1, S2 and S3, sessions 1, 2 and 3, respectively. * denotes significant differences ($p<0.05$).
