## Supplementary figure 3 for "Eccentric and Concentric Torque Feedback Training Induce Similar Clinical Improvements but Distinct Triceps Surae Motor Unit Adaptations in Non-Insertional Achilles Tendinopathy: A Randomized Controlled Trial"

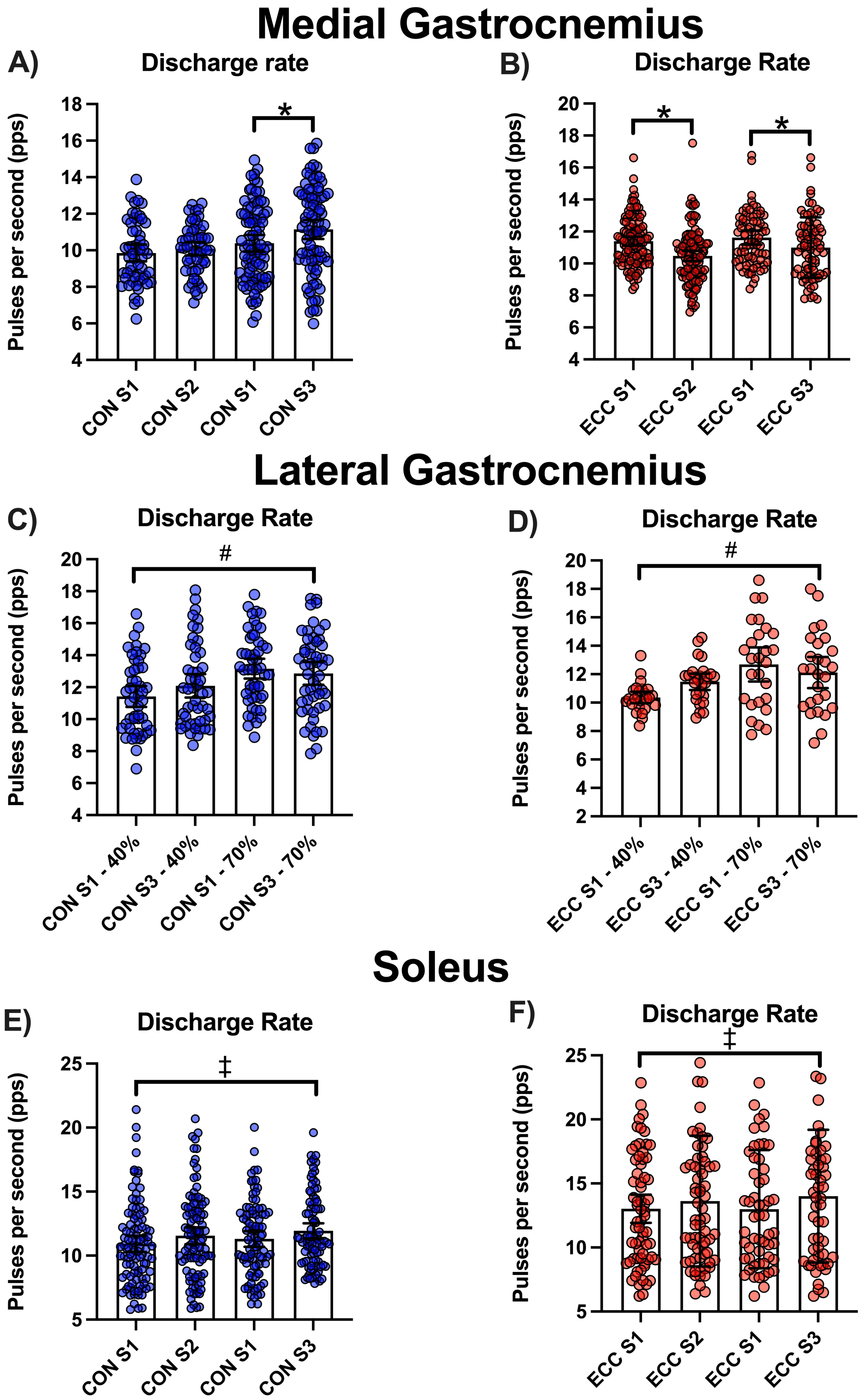


**Supplementary figure 3**. Changes in mean discharge rate for tracked motor units across the three sessions (S1, S2, S3) for concentric (CON) and eccentric (ECC) exercises. A) Mean discharge rate for gastrocnemius medialis CON. B) Mean discgarge rate for gastrocnemius medialis ECC. C) Mean discharge rate for gastrocnemius lateralis CON. D) Mean discharge rate for gastrocnemius lateralis ECC. E) Mean discharge rate for soleus CON. F) Mean discharge rate for soleus ECC. *, denotes significant differences for pairwise session comparisons ($p<0.05$). #, denotes a significant force x session interaction ($p<0.0001$). ‡, denotes a significant session effect ($p<0.05$).
